# Statin treatment effectiveness and the *SLCO1B1*\*5 reduced function genotype: long-term outcomes in women and men

**DOI:** 10.1101/2021.10.12.21264886

**Authors:** Deniz Türkmen, Jane A.H. Masoli, Chia-Ling Kuo, Jack Bowden, David Melzer, Luke C. Pilling

**Author notes:** **Corresponding author** Dr Luke Pilling, College House (room 1.09), Epidemiology and Public Health Group, University of Exeter, Magdalen road, St. Luke’s Campus, Exeter, EX1 2LU, UK.

## Abstract

**Objective:** To estimate the effect of the *SLCO1B1*\*5 genotype (decreases statin transport) on cholesterol control and treatment duration in male and female primary care patients prescribed common statin medications.

**Methods and Analysis:** 69,185 European-ancestry UK Biobank cohort participants prescribed simvastatin or atorvastatin (aged 40 to 79 years at first prescription; treatment duration 1 month to 29 years, mean 5.7 years). Principal outcomes were clinically high total cholesterol (>5mmol/L) at baseline, plus treatment discontinuation.

**Results:** 48.4% of 591 females homozygous for *SLCO1B1*\*5 decreased function genotype had raised cholesterol, vs. 41.7% of those with functioning SLCO1B1 (Odds Ratio 1.31: 95% Confidence Intervals 1.1 to 1.55, p=0.001). Fewer males had high cholesterol, and the genotype effect was attenuated. In primary care prescribing, females homozygous for *SLCO1B1*\*5 were more likely to stop receiving these statins (29.5%) than women with normal SLCO1B1 (25.7%) (Hazard Ratio 1.19: 95%CI 1.03 to 1.37, p=0.01), amounting to five discontinuations per 100 statin-years in the *SLCO1B1*\*5 group vs four in the normal SLCO1B1 function group. This remained significant after the first year of treatment (HR for discontinuing >1 year after first prescription 1.3: 95%CI 1.08 to 1.56; p=0.006). In men *SLCO1B1*\*5 was only associated with treatment discontinuation in the first year.

**Conclusions:** In this large community sample of patients on commonly prescribed statins, the *SLCO1B1*\*5 decreased function variant had much larger effects on cholesterol control and treatment duration in women than in men. Efforts to improve effectiveness of statin therapy in women may need to include *SLCO1B1*\*5 genotype-guided statin selection.

**What is already known about this subject:** Genetic variants affecting *SLCO1B1* (statin transporter) gene function increase concentrations of unmetabolized statin molecules (mostly simvastatin and atorvastatin). Previous studies of statin-treated patients have reported reduced likelihood of achieving target cholesterol levels plus increased adverse effects and medication non-adherence mainly in the first year of treatment.

However, little data have been available on key outcomes over longer follow-ups or on outcomes by sex, despite large differences in statin treatment patterns between men and women.

**What this study adds:** In 69,185 UK Biobank participants reporting simvastatin or atorvastatin use at baseline assessment, substantially more women had clinically high total cholesterol (>5 mmol/L) compared to men (42% vs. 25%). Female carriers of the *SLCO1B1*\*5 (decreased SLCO1B1 function) genetic variant were especially likely to have high cholesterol, despite being on statin treatment.

In primary care records of atorvastatin and simvastatin prescribing (>10 years follow-up), female carriers of *SLCO1B1*\*5 were more likely to stop statins. In men, *SLCO1B1*\*5 was only associated with discontinuing statin treatment in the first year after starting treatment.

## INTRODUCTION

Elevated low-density lipoprotein cholesterol (LDL-C) level is a major risk factor for myocardial infarction and stroke (1). Statins are the most commonly prescribed cholesterol-lowering drugs and reduce cardiovascular morbidity and mortality in higher risk patients (2,3). However a major barrier to effectiveness is medication non-adherence, often due to reported side effects including muscle pain (4). The STRENGTH study included 509 hypercholesterolaemic patients who were randomised to simvastatin, atorvastatin or pravastatin and discontinuation of treatment due to adverse effects was significant for simvastatin (Odds Ratio: 2.8, 95% Confidence Intervals 1.3–6.0) and atorvastatin (OR 1.6: 95%CI 0.7–3.7) in 16 weeks follow-up(5). However, a systematic review of randomised controlled trials (n=74 102) found no differences between placebo and statin groups for developing muscle symptoms and discontinuation of treatment (6), perhaps because trial participants are generally healthier than many older people prescribed statins.

Genetic polymorphisms in the solute carrier organic anion transporter 1B1 (*SLCO1B1*) gene, which encodes organic anion transporter polypeptide 1b1 (OATP1B1) and transports statins into tissues, may influence the effectiveness of lipid lowering therapy(7–11). Decreased hepatocellular concentrations of statins results in lower efficacy for reducing LDL-C, and increased systemic exposure to statins increases the risk of developing muscle weakness and muscle pain (12). *SLCO1B1*\*5 is a single nucleotide polymorphism (SNP, rs4149056 T>C) resulting in an amino acid substitution in SLCO1B1 (p.Val174Ala), increasing plasma levels of simvastatin by 221% and atorvastatin by 144% (13). A meta-analysis of 13 atorvastatin studies found *SLCO1B1*\*5 was associated with atorvastatin-related adverse drug reactions (OR 1.57, p=0.01) (14), yet literature linking *SLCO1B1*\*5 (5,9,10) to statin’s evidence is mixed and mostly focussed on shorter term outcomes (statin-related myotoxicity is predominantly reported in the first year of treatment, with median onset of 1 month after treatment initiation) (11). A 2013 study of UK primary care recruited 77 patients with statin-induced myopathy and found *SLCO1B1*\*5 significantly increased risk of myopathy compared to controls (OR per *5 allele 2.1, 95%CI 1.3 to 3.2) (15), but more research is needed linking genotype and GP data.

Though sex-differences in cholesterol levels are known, with LDL generally lower in men and higher total cholesterol levels in women whilst treated with statins (17), previous studies have mainly focused on men and effects in women are understudied (18–20). In addition, women have greater risk of adverse drug reactions, yet many cardiovascular risk models do not take into account female-specific factors (21). Historically UK guidelines for prescription of statins for cardiovascular disease (CVD) prevention used the same clinical cut points for high cholesterol (>5 mmol/L) and LDL (>3 mmol/L) for men and women(22). Current UK clinical recommendations are to begin atorvastatin treatment when 10-year CVD risk is >10% and to assess statin effectiveness if 40% reduction in non-HDL cholesterol is achieved 3 months after treatment initiation (23). Women have lower body weight and a higher percentage of body fat compared to men, which might lead to higher concentrations of lipophilic drugs such as simvastatin and atorvastatin (24) and increased risk of adverse events, which may be exacerbated by the *SLCO1B1*\*5 decreased function genotype which increases concentrations of unmetabolized drugs (12).

We therefore aimed to determine whether women prescribed simvastatin or atorvastatin were as likely as men to achieve cholesterol levels below clinically high cut points using data from the UK Biobank, a large cohort of community volunteers followed in primary care and hospital electronic medical records for over 10 years. Secondly, we tested whether the *SLCO1B1*\*5 decreased function genotype was associated with discontinuing statin treatment (in the first year and the longer term) in males and females separately. Statins are known to impact inflammation (25) and diabetes risk (3), so we also assessed C-reactive protein, alanine aminotransferase and HbA1c at baseline. We also investigated effects on self-reported side effects (including nausea and fatigue) and muscle symptoms. Considering the strong evidence for *SLCO1B1*\*5 affecting patients on simvastatin or atorvastatin and, that GPs prescribe them interchangeably, we examined both together.

## METHODS

### UK Biobank cohort

The UK Biobank recruited 503,325 community-based volunteers aged 40-70 years who visited one of 22 assessment centres in Wales, Scotland or England in 2006-2010 (26). Comprehensive questionnaires on demographic, lifestyle and health information data were collected at the baseline assessment. Blood samples for genetic and biochemical analyses, and anthropometric measures were taken. This study of atorvastatin and simvastatin comprises two distinct analysis sections. First using the data from baseline assessment only, and second using the linked GP (primary care) data available in 230,096 (45.7%) participants (Figure 1).

**Figure 1.**
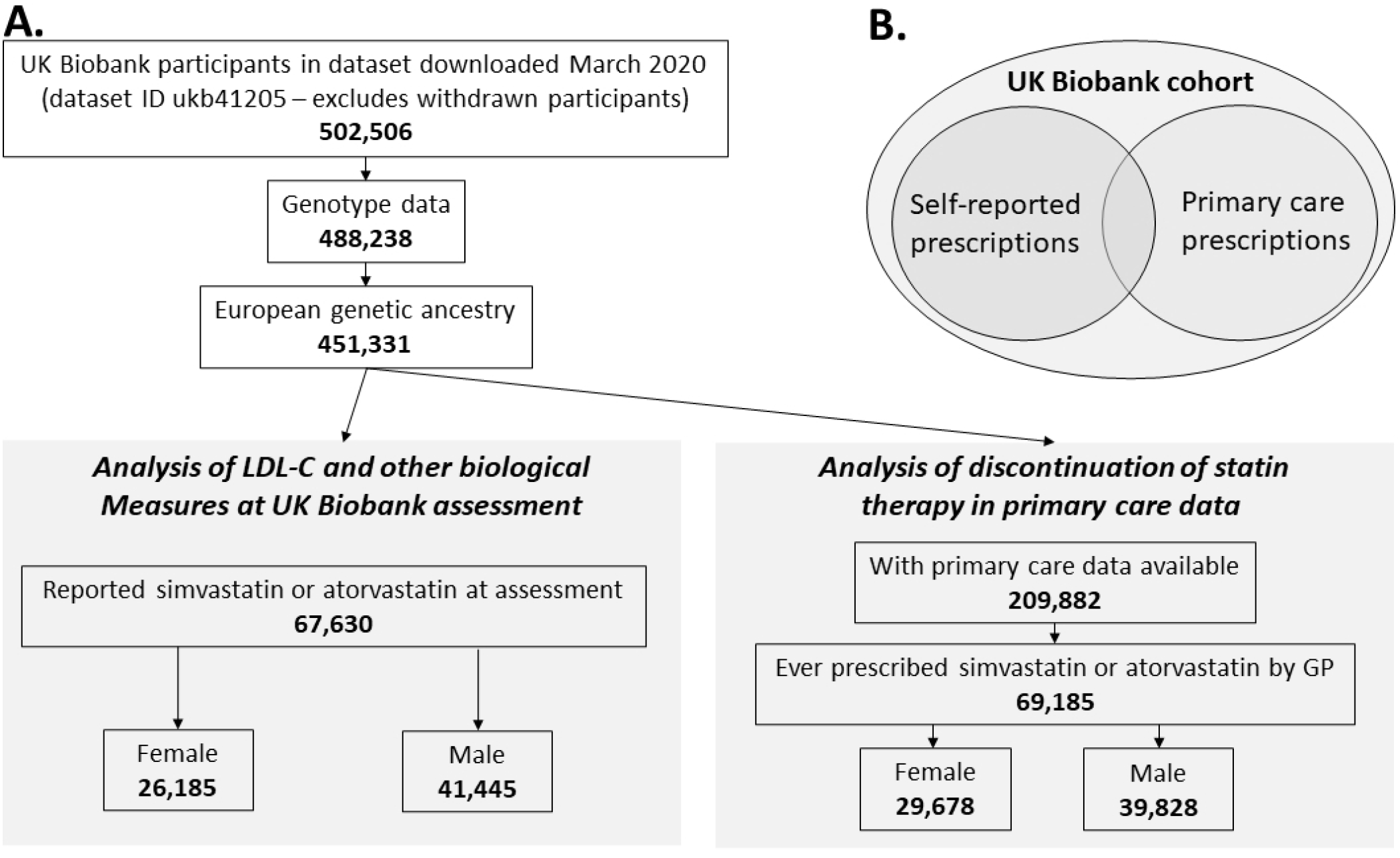
Cohort flowchart. A) shows a flowchart illustrating the number of UK Biobank participant eligible for analysis (i.e. with sufficient genetic and medication data). B) Illustrates the two subsets of UKB used in analyses: the baseline analysis of self-reported prescriptions (available in all participants), the primary care prescribing data available in ∼45% of the whole sample (up to 2017).

### Baseline assessment

The UK Biobank baseline assessment included self-reported medications; we analysed simvastatin and atorvastatin.

Lipid levels were measured in the UK Biobank baseline data and were categorized based on NHS reference levels at the time of assessment (22,27): high total cholesterol (>5mmol/L), high LDL-C (>3mmol/L), high triglycerides (>2.3mmol/L). We used pre-diabetic Hba1c level (>47mmol/mol) (28), high C-reactive protein (CRP) (>10mg/L) (29), high alanine aminotransferase (ALT) (>25 IU/L for females, >33 IU/L for males) (30) as statin use may worsen these variables.

We also analysed self-reported symptoms associated with statin use which may cause discontinuation of the treatment (31). We used “Headaches for 3+ months” (Data-Field: 3799), “Frequency of tiredness/lethargy in last 2 weeks” (Data field: 2080), and any reported pain for 3+ months (combining Data-Fields 3404, 3571, 3741, 3414, 3773, 2956).

### General Practice (GP) data

More than 57million prescriptions for 230,096 (45.7%) participants in the primary care data were available. The GP data was up to August 2016 (England TPP system supplier) and September 2017 (Wales EMIS/Vision system). Drug name, quantity, date of prescription, drug code (in clinical Read v2, British National Formulary (BNF), or dm+d (Dictionary of Medicines and Devices) format, depending on suppler) are available. We identified prescribing records for Simvastatin (Zocor, Simvastatin, Simvador) 10 mg, 20mg, 40mg and 80mg and for Atorvastatin (Atorvastatin, Lipitor) 10mg, 20mg, 40mg, 60mg and 80mg to analyse the date first prescribed, date last prescribed, number of total prescriptions, and average number of prescriptions over the treatment span.

Participants with a date of last prescription at least 3 months prior to the censoring date were defined as having discontinued treatment. The censoring date was either the date of deduction (removal from GP list, where available) or 28 Feb 2016 where no deduction date was present (i.e., still registered at an available practice). Data after 28 Feb 2016 is incomplete, depending on GP provider (see UK Biobank documentation (32)). We also evaluated prescriptions for other statins (Cerivastatin, Fluvastatin, Pravastatin and Rosuvastatin) to identify patients switching treatments from Simvastatin or Atorvastatin.

Muscle symptoms were ascertained from ICD-10 codes (33) and converted to Read codes used in UK primary care records (using UK Biobank-provided diagnostic code maps), available for up to 11 years follow-up after baseline assessment. We included ICD-10 codes for myopathy, myositis or myalgia (G72.0; G72.8; G72.9; M60.8; M60.9; M79.1).

### *SLCO1B1*5* genotype

Directly genotyped genetic variants (n=805,426) were obtained by UK Biobank using two near-identical platform: the Affymetrix Axiom UKB array (in 438,427 participants) and the Affymetrix UKBiLEVE array (in 49,950 participants). The central UK Biobank team performed imputation in 487,442 participants and the number of genetic variants reached ∼96 million (34). Different ancestral groups when analysed together can cause bias in genetic studies (35), thus we included 451,367 (93%) genetically European ancestry participants.

We analysed the *SLCO1B1*\*5 genetic variant rs4149056 (C allele, directly genotyped) with well-documented effects on simvastatin and atorvastatin related side effects in the literature, particularly on muscle symptoms (36). Genotype data was not returned to participants as part of the study.

### Statistical analyses

Associations between genotype and biochemical variables at the baseline were tested by logistic regression, adjusting for age and the first ten principal components of genetic ancestry to control for population substructure.

The association between genotype and discontinuation was tested using Cox’s proportional hazards regression models. We also performed Kaplan-Meier plots. Participants entered the model at the date of first prescription of statins and exited on the date of first incident outcome or end of records, thus providing an ‘intention to treat’ analysis reducing any effect of genetically associated discontinuation of treatment. We tested the associations between GP-diagnosed muscle symptoms that occurred in the first 3 months and 3 months or longer after the first prescription date using time to event models.

To estimate the *Genetically Moderated Treatment Effect* (GMTE) we used “TWIST” (Triangulation with a Study) (37): a novel pharmacogenetic causal inference approach to estimate population average effect on total cholesterol if all *SLCO1B1*\*5 homozygotes could experience the same treatment effect as non-carriers. In brief, several assumptions common to pharmacogenetic analysis are tested (primarily that genetic variant *SLCO1B1*\*5: does not predict whether an individual receives statin treatment; is not associated with any confounders predicting statin use or cholesterol; and only affects cholesterol through the interaction with statins). From this analysis the most efficient and robust estimate of the GMTE is derived.

### Sensitivity analysis

a. We conducted competing risk regression model for discontinuation or death to check whether the estimate is drastically changed when accounting for the competing risk of mortality.
b. We also tested for interactions between *SLCO1B1* genotype and sex with discontinuation of treatment within year 1 and discontinuation after one year of treatment.

### Patient and public involvement

Patients and participants are extensively involved in the UK Biobank study itself. No patients were involved in developing the research question or the outcomes tested in this analysis.

## RESULTS

### Characteristics and associations at UK Baseline assessment

There were 26,185 female and 41,445 male European-ancestry UK Biobank participants who reported atorvastatin or simvastatin treatment at baseline assessment. The mean age was 61.6 years (SD 5.7) for females and 61.4 (SD 6.1) for males (Table 1) (see Supplementary Table 1 for details including heterozygotes).

**Table 1.** Characteristics of UK Biobank participants on simvastatin or atorvastatin therapy.

|  |  | *SLCO1B1* *5 status<sup>∞</sup> |  |  |  |
| --- | --- | --- | --- | --- | --- |
|  |  | Female |  | Male |  |
|  |  | Normal function (*1/*1) | Reduced function (*5/*5) | Normal function (*1/*1) | Reduced function (*5/*5) |
| **- Baseline assessment (self-reported)** |  |  |  |  |  |
| n (% of 26,185 females or 41,445 males) |  | 18,925(72.27) | 591(2.26) | 29,996(72.38) | 927(2.24) |
| Age, years | Min-max | 40-70 | 41-70 | 40-70 | 40-70 |
|  | Mean (SD) | 61.7(5.7) | 61.6(5.8) | 61.4(6.1) | 61.6(6.1) |
| Weight, kg | Mean (SD) | 76.3 (15.4) | 76.3 (15.7) | 89.3(15.2) | 89.8(14.9) |
| BMI | Mean (SD) | 29.5(5.6) | 29.5(5.6) | 29.31(4.5) | 29.4(4.3) |
| LDL, n >3mmol/L (% of genotype group) |  | 6,492(36.17) | 250(44.8) | 7,743(27.1) | 267(30.2) |
| Triglycerides, n >2.3mmol/L (%) |  | 5,040(26.63) | 178(30.12) | 9,841(32.81) | 345(37.22) |
| Total cholesterol, n >5mmol/L (%) |  | 7,485(41.65) | 270(48.39) | 7,069(24.7) | 258(29.05) |
| HbA1c, n >47mmol/mol (%) |  | 2,526(13.99) | 105(18.52) | 4,558(15.94) | 124(13.98) |
| **- Primary Care data** |  |  |  |  |  |
| n (% of 29,574 females or 39,611 males) |  | 21,345(72.17) | 691(2.34) | 28,608(72.22) | 947(2.39) |
| Age at first statin prescription | Min-max | 40-78.9 | 40.3-77.31 | 40-79.2 | 41.1-78.2 |
|  | Mean (SD) | 61.9 (7.1) | 61.9(7.3) | 60.9 (7.2) | 61.1(6.9) |
| Years between first and last statin* | Min-max | 0.002-28.2 | 0.01-24.7 | 0.002-27.2 | 0.01-29.3 |
|  | Mean (SD) | 5.7(4.8) | 5.4(4.5) | 6.6 (4.8) | 6.6(4.9) |
| Muscle diagnoses^ prior to statin* | n (%) | 560(2.62) | 19(2.75) | 499(1.74) | 19(2.01) |
| MI/angina diagnoses¥ prior to statin* | n (%) | 1,078(5.05) | 36(5.21) | 3,151(11.01) | 107(11.3) |
| Muscle diagnoses after first statin* | n (%) | 776(3.64) | 26(3.76) | 880(3.08) | 34(3.59) |
| MI/angina after first statin* | n (%) | 2,875(13.47) | 98(14.18) | 6,978(24.39) | 211(22.28) |
| Discontinuation ever, n (%) | n (%) | 5,476(25.65) | 204(29.52) | 6,119(21.39) | 212(22.39) |
| Discontinuation in 1 year, n (%) | n (%) | 2,489(11.66) | 86(12.45) | 2,333(8.15) | 95(10.03) |
| Discontinuation in year 1+, n (%) | n (%) | 2,987(17.62) | 118(21.77) | 3,786(15.49) | 117(14.53) |

<sup>∞</sup>rs4149056 genotype ("Reduced function" = CC homozygotes; "Normal function" = TT homozygotes), ^Primary care-diagnosed muscle symptoms (myopathy, myositis or myalgia), ¥Hospital inpatient diagnosis of MI or angina, \*simvastatin or atorvastatin prescription. See Supplementary Table 1 for full table including heterozygotes (\*1/\*5 group).

42.1% (10,485/24,907) of women and 25.3 % (9,995/39,527) of men had clinically high total cholesterol levels (>5mmol/L), significant in logistic regression models adjusted for age (Odds Ratio 2.2: 95% Confidence Intervals 2.11 to 2.26, p=5*10^−439^). The association was significant and effect consistent after further adjustment for assessment centre, highest education level attained, weight, waist circumference, and smoking status (OR 1.94: 95%CI 1.86 to 2.01, p=8*10^−256^).

The *SLCO1B1*\*5 impaired statin intracellular transport genotype (rs4149056 CC homozygous) was present in 2.26% of female study participants (n=591). This group was more likely to have clinically raised total cholesterol compared to females with normal function (rs4149056 TT homozygous) genotype (OR 1.31: 95%CI 1.10 to 1.55, p=0.001) in logistic regression models adjusted for age and genetic principal components of ancestry 1 to 10 (Table 2). 48.4% of female *SLCO1B1*\*5 homozygotes had raised cholesterol, compared to 41.7% of the *SLCO1B1* normal function group (excess 6.7%: 95%CI 2.6 to 10.9, p=0.001). Females *SLCO1B1*\*5 homozygotes were also more likely to have raised LDL (OR 1.42: 95%CI 1.2 to 1.69, p=4.4*10^−5^) (44.8% vs. 36.2%; excess 8.6%: 95%CI 4.3 to 12.6, p=4*10^−5^).

**Table 2.**
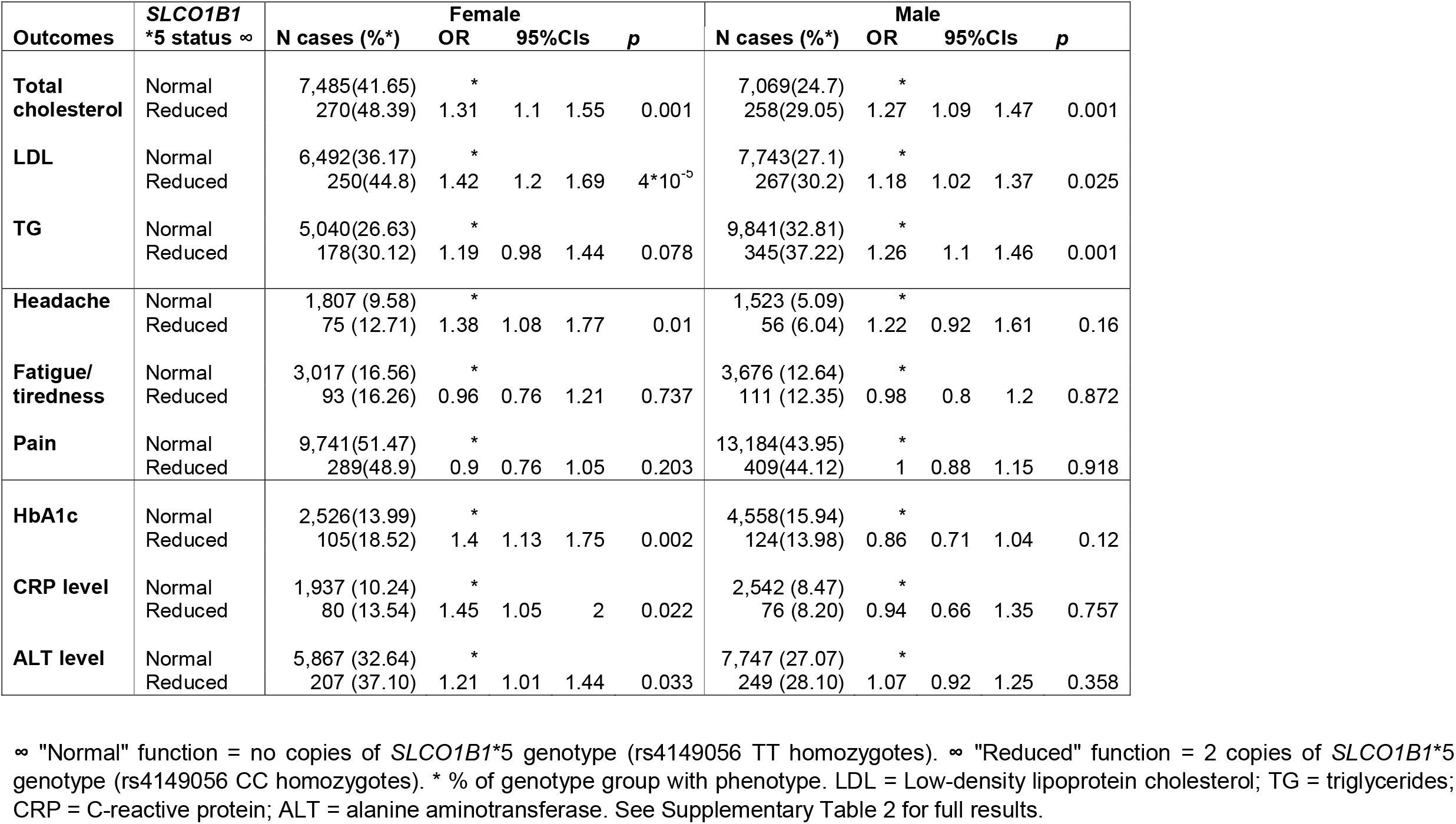
SLCO1B1 genotype associations with baseline analyses in patients who reported statin treatment.

In our TWIST causal analysis (37) of high/low total cholesterol we estimated that if all female *SLCO1B1*5* homozygotes could experience the same treatment effect as non-carriers their risk of high cholesterol would reduce by 6.34% (95%CI 3.33 to 9.35, p=3.7*10^−5^) i.e. from 48.4% (the number of female *SLCO1B1*\*5 homozygotes with high cholesterol) to 42.1%. This equates to 37 women (6.34% of 591 female *SLCO1B1*5* homozygotes in analysis). In a complimentary analysis treating total cholesterol as a continuous outcome we estimated that *SLCO1B1*\*5 homozygous females would have 0.147 mmol/L lower total cholesterol if they could be treated with a lipid-lowering medication unaffected by the genotype. This suggested a reduction in the number of *5 homozygotes with high total cholesterol from 48.4% to 40.7%, corresponding to 46 *SLCO1B1*\*5 homozygous women with cholesterol <5mmol/L where currently it is >5mmol/L. We used the ‘robust’ GMTE estimate, which estimates the GMTE in treated individuals, then subtracts the GMTE estimate in untreated (but eligible) individuals. This guards against off-target genetic effects that could directly influence the likelihood of being treated with a statin and/or an individual’s cholesterol level.

Males homozygous for the *SLCO1B1*\*5 decreased function variant (n=927, 2.24% of 41,445 males in study) were also more likely to have raised total cholesterol than *SLCO1B1* normal function homozygotes (29.1% vs. 24.7%; OR 1.27: 95%CI 1.09 to 1.47, p=0.001), with similar trends for raised LDL (30.2% vs. 27.1%; OR 1.18: 95%CI 1.02 to 1.37, p=0.025) (Table 2).

At UK Biobank baseline, female *SLCO1B1*\*5 homozygotes were more likely to report having headaches for ≥3 months than women homozygous for the *SLCO1B1* normal function genotype (OR 1.58: 95%CI 1.1 to 2.29; p=0.01), but there was no association with frequency of tiredness / lethargy in last 2 weeks and chronic pain for ≥3 months (Table 2). High CRP level, ALT level, Hb1Ac and total cholesterol was associated with *SLCO1B1*\*5 homozygous genotype in females (Table 2). Males homozygous for *SLCO1B1*5* were not more likely to report headaches, fatigue or pain compared to normal function homozygotes.

### GP prescribing data on simvastatin and atorvastatin

There were 29,574 female and 39,611 male UK Biobank participants of European ancestry who received at least one prescription of simvastatin or atorvastatin in the available GP data (from 1990 to 2017, see Methods and Figure 1). The length of simvastatin or atorvastatin treatment spanned from a single prescription to 607 prescriptions over 28.2 years (mean 5.7 years, SD 4.7 in females).

Female *SLCO1B1*\*5 homozygotes (n=691) ranged from a single prescription to 24.7 years (mean 5.4, SD 4.5) whereas in male (n=947) prescriptions were up to 29.3 years (mean 6.6, SD 4.9). See Table 1 for details.

### *SLCO1B1*\*5 association with discontinuing simvastatin or atorvastatin treatment

We identified patients who discontinued atorvastatin or simvastatin treatment as those where their last prescription date was >3 months prior to the censoring date of the GP data (or death). Participants with prescriptions on or after the censoring date are assumed to have not discontinued. The overall rate of discontinuation was 23.3% (16,139 of 69,185 participants included in the analysis) i.e. of 69,185 participants who received at least one prescription of simvastatin or atorvastatin in the available GP data 53,046 (76.7%) were still on treatment.

29.5% of female *SLCO1B1*5* homozygotes discontinued vs 25.6% of normal function (Table 1). The association was significant in time-to-event analysis adjusted for age and genetic principal component of ancestry (HR: 1.19, 95%CI 1.03 to 1.37, p=0.015). Yet male *SLCO1B1*5* homozygotes were not more likely to discontinue (HR 1.05: 95%CI 0.92 to 1.2, p=0.44). See Figure 2A for detailed estimates and Figure 2B for cumulative incidence plots of *SLCO1B1*\*5 homozygotes association with discontinuing treatment (plots are truncated to 15 years for clarity; full plot Supplementary Figure 1; see Supplementary Table 2 for details). There was a significant interaction between sex and *SLCO1B1*\*5 with discontinuation of treatment (p=0.02). The association between genotype and discontinuation in females was consistent in Fine and Gray’s competing risks models accounting for the competing risk of mortality (sub-HR: 1.19, 95%CI 1.04 to 1.37, p=0.012).

**Figure 2.**
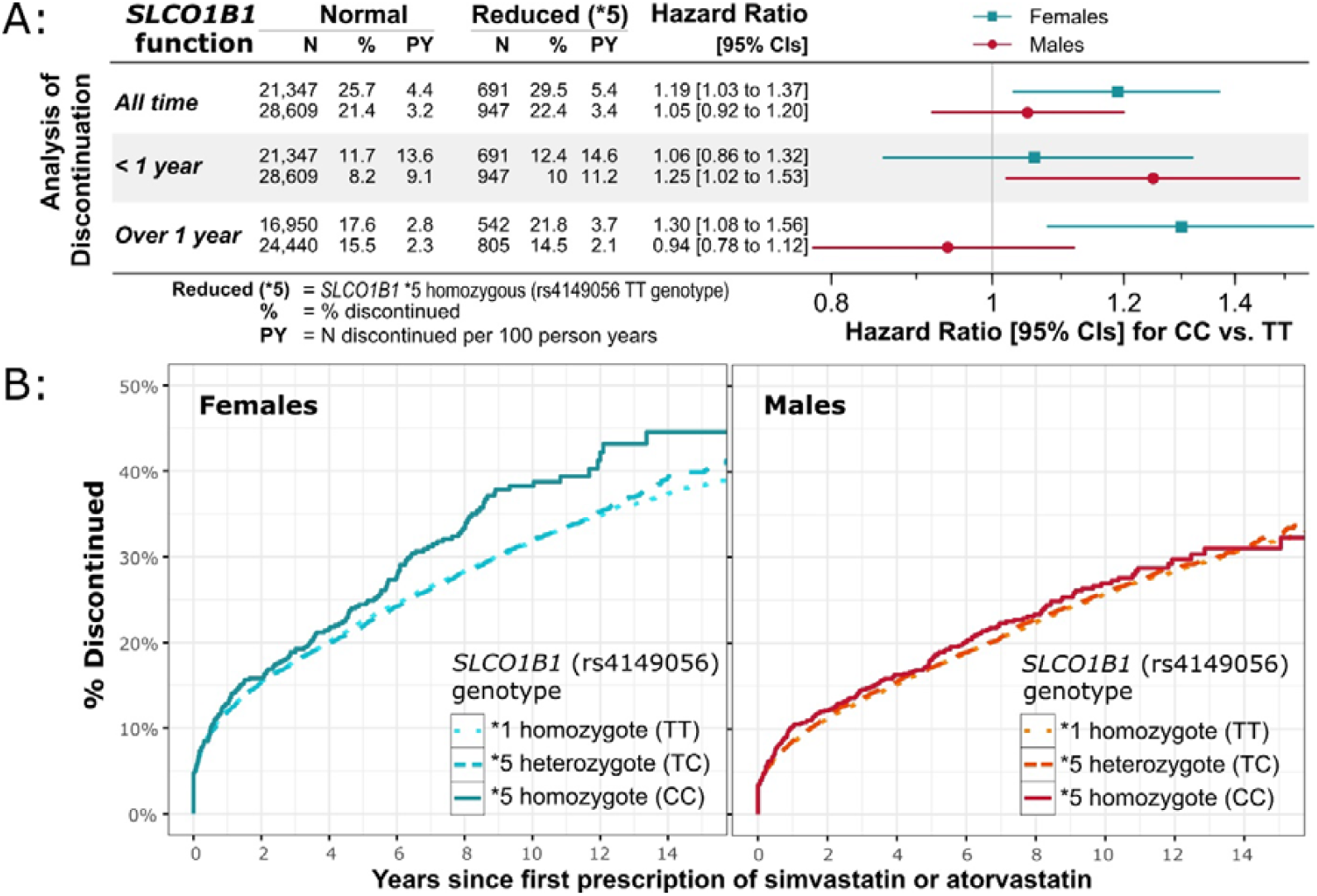
*SLCO1B1*5* genotype association with discontinuing GP-prescribed simvastatin and atorvastatin treatment. Associations between *SLCO1B1*5* genotype (reduced function compared to normal genotype, i.e. rs4149056 CC homozygotes vs TT homozygotes) and discontinuing GP-prescribed simvastatin or atorvastatin treatment in males and females separately. 2A) shows the number of “cases” (discontinuing treatment) and “controls” (remained on treatment) for the normal and reduced-function homozygous groups, the number of discontinuations per 100 person years on treatment in the two groups, and the Hazard Ratio from Cox’s proportional hazards regression models. Also shown are the associations from stratified analyses of short term (stopped less than one year after beginning treatment) and longer term (stopped greater than one year after beginning treatment). See Supplementary Table 2 for details, including for the normal/reduced (*1/*5) heterozygous group. 2B) show the cumulative incidence over time of discontinuing treatment in males and females, stratified by *SLCO1B1*5* genotype. X-axis is censored at 15 years for figure clarity, see Supplementary Figure 1 for complete plots.

In analysis stratified by whether discontinuation occurred within 12 months of beginning treatment or greater than 12 months, male decreased function homozygotes were more likely to discontinue in the first 12 months (HR 1.25, 95%CI 1.02 to 1.53, p=0.03), whereas female decreased function homozygotes were more likely to discontinue treatment in the long term (1+ years) (HR 1.3, 95%CI 1.08 to 1.56, p=0.006) (Figure 2A).

2,160 women who discontinued treatment of Simvastatin or Atorvastatin switched to another statin within 12 months of discontinuation (1208 to Pravastatin, 895 to Rosuvatatin, 56 to Fluvastatin and 1 to Cerivastatin). 24.5% of female *SLCO1B1*5* homozygotes who discontinued Simvastatin or Atorvastatin switched treatment compared to 24.3% of normal function carriers (OR 1.04: 95%CI 0.75 to 1.44, p=0.81). See Supplementary Table 3 for all the details.

In secondary analysis we investigated whether sufficient data on dose of atorvastatin or simvastatin in *SLCO1B1*5* homozygotes at the time of discontinuation was available, though this analysis is limited by the low number of homozygotes in different dose groups and due to lack of data on instructions from GPs (e.g. number of tablets per day), data which is not available in the UK Biobank-linked GP data. See Supplementary Table 4 for tabulation of available data.

### *SLCO1B1*\*5 associations with GP-diagnosed muscle symptoms

110 female and 96 male participants had GP recorded muscle symptoms in the 3 months after their first prescription of atorvastatin or simvastatin. 848 female and 1026 male participants had GP recorded muscle symptoms greater than 3 months after the first prescription of atorvastatin or simvastatin (i.e. the stable treatment period). This was lower than expected based on previous literature (2,5,9), with similar rates in the different genotype groups. In the first 3 months, there was no significant association between muscle symptoms and genotype. After 3 months, female *SLCO1B1*\*5 heterozygotes were more likely to experience muscle symptoms compared to female normal function homozygotes (OR 1.19, 95%CI 1.03 to 1.4, p=0.02). However, there was no significant association with muscle symptoms in males in any group of *SLCO1B1*\*5 genotype (see Supplementary Table 5).

## DISCUSSION

We aimed to assess the success of cholesterol control in men and women taking simvastatin or atorvastatin, including examining the contribution of the *SLCO1B1*\*5 genetic variant that impairs intracellular transport of these statins. In UK Biobank participants taking simvastatin or atorvastatin at baseline assessment, 42% of females and 25% of males had clinically high total cholesterol (>5mmol/L), despite treatment (OR 2.2: 95%CI 2.11 to 2.26, p=5*10^−439^), with consistent results for LDL also observed. We found that females homozygous for *SLCO1B1*\*5 (i.e. with reduced protein function) were more likely to have high cholesterol (compared to common “normal function” homozygotes). In the linked GP electronic clinical records data, female *SLCO1B1*\*5 reduced function homozygotes were more likely to discontinue simvastatin or atorvastatin treatment: 5 discontinuations per 100 patient years on statins, compared to 4 per 100 in the normal function genotype group.

The difference between males and females could be due in part to males being more likely to adhere to statin therapy (38), and is consistent with previous reports of females having higher total cholesterol levels whilst treated with statins (17). UK guidelines for prescription of statins for prevention of CVD at the time of UK Biobank baseline assessment used the same clinical cut point for high total cholesterol (>5 mmol/L) in males and females (22). The current UK clinical recommendations are to assess statin effectiveness by measuring percentage reduction in non-HDL cholesterol after 3 months of treatment (23), in part acknowledging the sex difference (this analysis was not possible in the cross-sectional UK Biobank cholesterol data). A systematic review of randomised controlled trials (RCTs) with 74,102 subjects found that statin therapy was not associated with discontinuation of treatment compared with placebo (10), yet our analysis shows that for a subset of patients – especially females carrying the *SLCO1B1*\*5 genotype – medication adherence is affected.

As the studied statins are mainstays of cardiovascular disease prevention, and non-adherence is a major barrier to treatment effectiveness, prescribing an appropriate statin without high risk of adverse events and with higher efficacy at first intervention could reduce discontinuation and improve control of cholesterol, especially in women. We used TWIST (37): a novel pharmacogenetic causal inference framework for estimating population average genetically modified treatment effect (GMTE) on total cholesterol if all *SLCO1B1*\*5 homozygotes could experience the same treatment effect as non-carriers. Treating total cholesterol as a binary outcome (>5mmol/L vs. <5mmol/L) we estimate their risk of high total cholesterol would reduce 6.34% from 48.4% to 42.1%. Of the 591 female *SLCO1B1*\*5 homozygotes in the study, this corresponds to 37 women meeting the cholesterol target if they could be prescribed a lipid-lowering medication not affected by *SLCO1B1*\*5. Next, we repeated the analysis treating total cholesterol as a continuous outcome. From this we estimated that female *5 homozygotes would have 0.147 mmol/L lower total cholesterol if they could be treated with a lipid-lowering medication unaffected by the genotype. This suggested a reduction in the number of *5 homozygotes with high total cholesterol from 48.4% to 40.7%, corresponding to 46 *SLCO1B1*\*5 homozygous women who currently have high total cholesterol (>5mmol/L) gaining control if prescribed an alternative lipid-lowering medication not affected by *SLCO1B1*\*5.

In males we found no excess discontinuation in *SLCO1B1*\*5 homozygotes when analysing the whole prescribing period (3 discontinuations per 100 statin-years in both *5 homozygotes and “normal” functioners). Previous analyses have specifically investigated the first 12 months after beginning statin therapy (41,42), and in analysis restricted to this period male *5 homozygotes were more likely to discontinue than the normal function homozygote group, whereas females were more likely to discontinue in the longer term. The difference between males and females in both cholesterol control whilst on simvastatin or atorvastatin treatment, and in likelihood of discontinuing treatment, may have implications for interventions (specific statin prescribed, and dose) and subsequent cardiovascular outcomes. This significant interaction between *SLCO1B1*\*5 genotype and sex could be due to females having lower mean muscle mass and body weight, and higher percentage of body fat compared to males, leading to higher concentrations of unmetabolized simvastatin and atorvastatin (24), with *SLCO1B1*\*5 therefore causing increased discontinuation of treatment due to side effects (12).

Though we observe raised cholesterol levels in *SLCO1B1*\*5 homozygotes at the UK Biobank baseline assessment, and increased likelihood of discontinuing GP-prescribed simvastatin and atorvastatin therapy, we found limited evidence of *SLCO1B1*\*5 associations with GP-diagnosed muscle symptoms. This could be due to underreporting of statin-associated pain (by the patients themselves, or under-recording by GPs in the clinical record; 3.4% of participants prescribed simvastatin or atorvastatin received a relevant GP diagnosis (including myalgia, myositis), compared to previous studies with systematic ascertainment of muscle effects that reported up to 25% of patients with muscle symptoms (5,9,43). A recent systematic review of RCTs reported that statins were not associated with clinically confirmed muscle disorders, consistent with our results (patients may report muscle symptoms, but these are not clinically confirmed) (44). Additionally, our ability to analyse dose was limited due to data availability, and *SLCO1B1*\*5 has been linked to muscular complaints (and atorvastatin intolerance) especially in patients receiving high doses of atorvastatin (45). However, statins are known to impact inflammation (25) and diabetes risk (3): we found that female *SLCO1B1*\*5 homozygotes (but not male) treated with atorvastatin or simvastatin at baseline assessment had higher C-reactive protein, alanine aminotransferase and HbA1c, further emphasising the increased importance of appropriate prescribing in females.

Additionally, it is thought that a “nocebo” effect is common, where patients treated with statins report more symptoms thought to be associated with statins (4). Yet we find genotype (which the participants and GPs were not told about) to be associated with both cholesterol and discontinuation, so it is likely that statin pharmacokinetics and pharmacodynamics are being affected and causing adverse effects: as genotypes are inherited at conception and are not altered by later factors, associations with genotypes provide less confounded evidence than conventional observational associations (46). This is supported by our findings that female UK Biobank *SLCO1B1*\*5 homozygotes who self-reported statin therapy at baseline were significantly more likely to report headaches than normal function homozygotes (12.7% of *5 group reported chronic headache for 3+ months compared to 9.6% of normal group). However, we did not find a difference in reports of chronic pain (pain in any site for 3+ months), perhaps because the available questions in UK Biobank did not ask about muscle pain specifically.

This study has several strengths: it is a large cohort study with longitudinal electronic primary care and hospital medical records follow-up of patients, with a follow-up period considerably longer than in most previous studies, combined with genotype and self-report data. Yet there are several limitations, firstly that the UK Biobank participants are somewhat healthier than the general population (47). The magnitude of poor control of cholesterol in women in this relatively healthy cohort is therefore particularly disappointing. Secondly, only 45% of participants have available primary care data at the time of analysis – when more data becomes available, further investigation of prescriptions and diagnoses will be possible. As noted, questions about pain at UK Biobank baseline were nonspecific, and recording of statin adverse effect related symptoms in medical records may not be complete. However, given that study participants were not informed about their genotypes, the associations observed for metabolizer status are likely robust. Future work could improve ascertainment of adverse effects with systematic measurements, including of biochemical evidence of muscle damage. A further limitation is that we have no direct data on why the studied women were not moved onto different statins or prescribed sufficient doses to achieve target cholesterol levels.

Trials are showing that genotype-guided treatment for antiplatelet therapy reduces adverse events (48). Though a recent systematic review reported that safety concerns were limited in statin therapy (44) this was in the general population; in sub-populations carrying the *SLCO1B1*\*5 decreased function variant, especially women, tailoring the specific statin or dose may improve outcomes, and could highlight patients to target with novel agents for cholesterol lowering (49).

In conclusion, in the large UK Biobank community volunteer study, women prescribed atorvastatin or simvastatin were more likely to still have clinically raised cholesterol levels than men. In women, the *SLCO1B1*\*5 decreased function variant was associated with raised cholesterol levels and discontinuation of treatment during a follow-up of >10 years. Efforts are needed to improve effectiveness of statin therapy in women, including establishing whether *SLCO1B1*\*5 genotype-guided statin selection to one without high risk of adverse events and with higher efficacy in that patient could aid in reducing discontinuation rates.

## Data Availability

All data produced in the present study are available upon reasonable request to the authors

## Acknowledgements

Access to UK Biobank resource was granted under Application Number 14631. We would like to thank UK Biobank participants and coordinators for this dataset. The authors would like to acknowledge the use of the University of Exeter High-Performance Computing (HPC) facility in carrying out this work.

## Contributors

DT performed analyses, interpreted results, created the figures, searched literature, and co-wrote the manuscript. JM provided expert clinical interpretation of the data and contributed to the manuscript. CK and JB contributed to data analysis and interpretation, and contributed to the manuscript. DM oversaw interpretation and literature searching, and co-wrote the manuscript. LP generated data, performed analyses, interpreted results, created the figures, searched literature, and co-wrote the manuscript. LP is the guarantor. The corresponding author attests that all listed authors meet authorship criteria and that no others meeting the criteria have been omitted.

## Ethical Approval

The North West Multi-Centre Research Ethics Committee approved the collection and use of UK Biobank data (Research Ethics Committee reference 11/NW/0382).

## Data Availability Statement

The genetic and phenotypic UK Biobank data are available upon application to the UK Biobank (www.ukbiobank.ac.uk/register-apply). The derived data fields used in our analysis will be available via the UK Biobank, searching for application number 14631 - we are not able to share these directly.

