## Supplementary for "Statin treatment effectiveness and the *SLCO1B1*\*5 reduced function genotype: long-term outcomes in women and men"

Türkmen *et al.* 2021

**Supplementary Information**

### Supplementary Figure 1

Uncensored Kaplan-Meier plots for effect of rs4149056 C genotype (*SLCO1B1* *5) on discontinuation of atorvastatin and simvastatin in UK Biobank primary care data

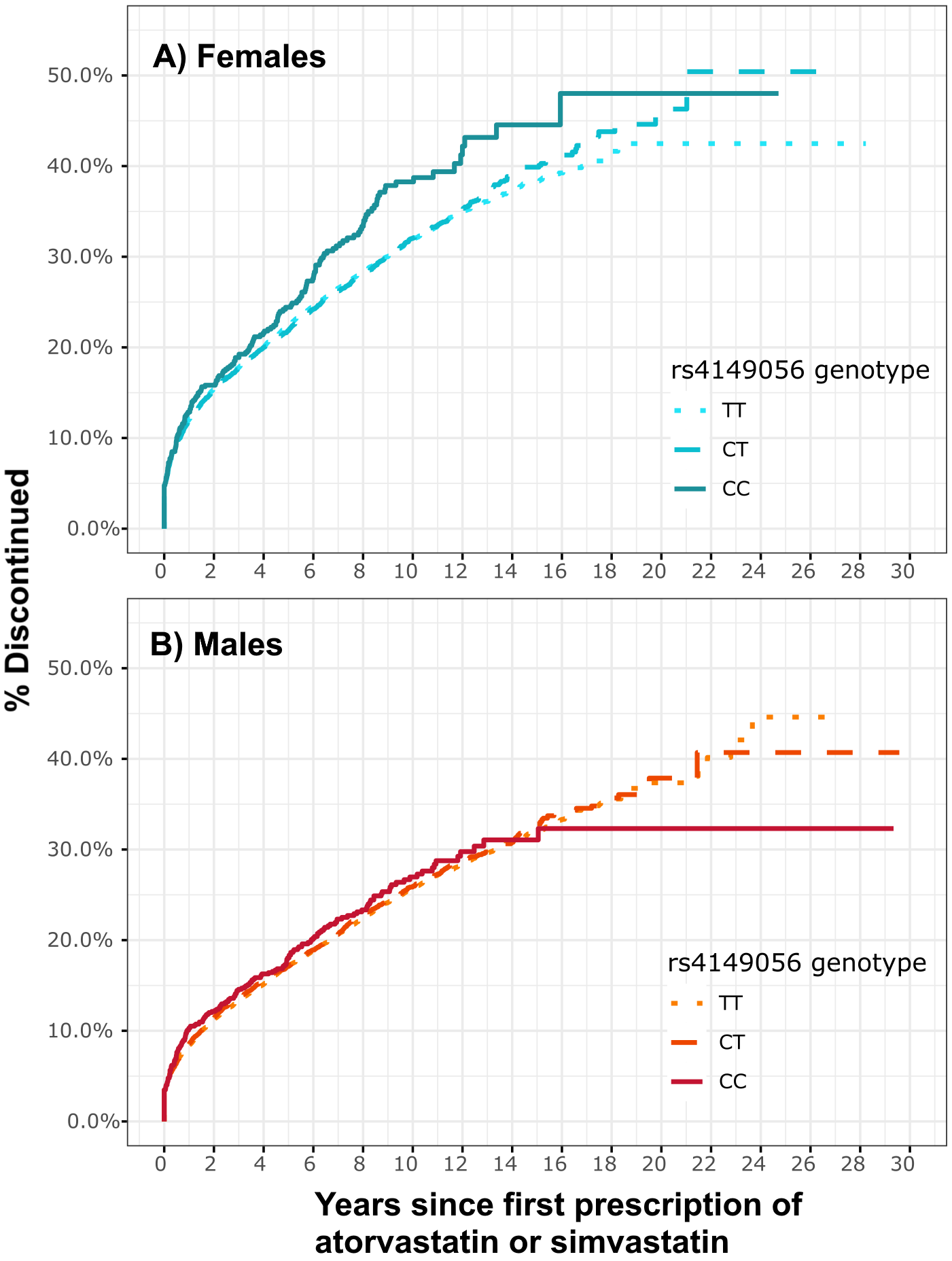

Genetic variant rs4149056 (*SLCO1B1* *5) C-allele associations with discontinuing GP-prescribed simvastatin or atorvastatin treatment in males and females separately. Plots show the cumulative incidence over time of discontinuing treatment in females (A) and males (B), stratified by rs4149056 genotype.

### Supplementary Table 1

Expanded descriptive summary statistics table for UK Biobank participants included in analysis

|  |  | **rs4149056 (*SLCO1B1* *5) genotype** | | | | | |
| --- | --- | --- | --- | --- | --- | --- | --- |
|  |  | **Female** | | | **Male** | | |
|  |  | **TT (*1/*1) homozgotes** | **CT (*1/*5) heterozygotes** | **CC (*5/*5) homozygotes** | **TT (*1/*1) homozgotes** | **CT (*1/*5) heterozygotes** | **CC (*5/*5) homozygotes** |
| ***- Baseline assessment (self-reported)*** | |  |  |  |  |  |  |
| n (% of genotype group) |  | 18,925(72.27) | 6,669(25.47) | 591(2.26) | 29,996(72.38) | 10,522(25.39) | 927(2.24) |
| Age | Min-max | 40-70 | 40-70 | 41-70 | 40-70 | 40-70 | 40-70 |
|  | Mean (SD) | 61.7(5.7) | 61.5(5.7) | 61.6(5.8) | 61.4(6.1) | 61.3(6) | 61.6(6.1) |
| Weight | Mean (SD) | 76.3 (15.4) | 76.2 (15.2) | 76.3 (15.7) | 89.3(15.2) | 89.2(15.0) | 89.8(14.9) |
| BMI | Mean (SD) | 29.5(5.6) | 29.4(5.6) | 29.5(5.6) | 29.31(4.5) | 29.3(4.4) | 29.4(4.3) |
| LDL, n >3mmol/L (% of genotype group) | | 6,492(36.17) | 2,367(37.18) | 250(44.8) | 7,743(27.1) | 2,845(28.46) | 267(30.2)) |
| Triglycerides, n >2.3mmol/L (%) | | 5,040(26.63) | 1,831(27.46) | 178(30.12) | 9,841(32.81) | 3,660(34.78) | 345(37.22) |
| Total cholesterol, n >5mmol/L (%) | | 7,485(41.65) | 2,730(42.80) | 270(48.39) | 7,069(24.7) | 2,668(26.64) | 258(29.05) |
| HbA1c, n >47mmol/mol (%) | | 2,526(13.99) | 889(14) | 105(18.52) | 4,558(15.94) | 1,572(15.74) | 124(13.98) |
| ***- Primary Care data*** |  |  |  |  |  | | |
| n (% of genotype group) |  | 21,345(72.17) | 7,538(25.49) | 691(2.34) | 28,608(72.22) | 10,056(25.39) | 947(2.39) |
| Age at first statin prescription | Min-max | 40-78.9 | 40-79.1 | 40.3-77.31 | 40-79.2 | 40-79.1 | 41.1-78.2 |
|  | Mean (SD) | 61,9 (7.1) | 61.8(7.1) | 61.9(7.3) | 60.9 (7.2) | 60.8(7.2) | 61.1(6.9) |
| Years between first and last statin* | Min-max | 0.002-28.2 | 0.002-26.3 | 0.01-24.7 | 0.002-27.2 | 0.002-29.5 | 0.01-29.3 |
|  | Mean (SD) | 5.7(4.8) | 5.7(4.7) | 5.4(4.5) | 6.6 (4.8) | 6.6(4.9) | 6.6(4.9) |
| Muscle diagnoses prior to statin* | n (%) | 560(2.62) | 190(2.52) | 19(2.75) | 499(1.74) | 154(1.53) | 19(2.01) |
| MI/angina diagnoses prior to statin* | n (%) | 1,078(5.05) | 400(5.31) | 36(5.21) | 3,151(11.01) | 1,158(11.52) | 107(11.3) |
| Muscle diagnoses after first statin* | n (%) | 776(3.64) | 312(4.14) | 26(3.76) | 880(3.08) | 314(3.12) | 34(3.59) |
| MI/angina after first statin* | n (%) | 2,875(13.47) | 995(13.20) | 98(14.18) | 6,978(24.39) | 2,431(24.17) | 211(22.28) |
| Discontinuation ever, n (%) | n (%) | 5,476(25.65) | 1,946(25.82) | 204(29.52) | 6,119(21.39) | 2,182(21.7) | 212(22.39) |
| Discontinuation in 1 year, n (%) | n (%) | 2,489(11.66) | 865(11.48) | 86(12.45) | 2,333(8.15) | 842(8.37) | 95(10.03) |
| Discontinuation in year 1+, n (%) | n (%) | 2,987(17.62) | 1,081(18.09) | 118(21.77) | 3,786(15.49) | 1,340(15.59) | 117(14.53) |

* simvastatin or atorvastatin prescription

### Supplementary Table 2

*SLCO1B1* *5 genotype association with discontinuing simvastatin and atorvastatin treatment in the GP prescribing data

|  | **Sex** | ***SLCO1B1* genotype** | **N** | **N disc*** | **Person-years** | **Discontinuations per 100 statin-years** | **HR** | **95% CIs** | | ***p*** |
| --- | --- | --- | --- | --- | --- | --- | --- | --- | --- | --- |
| Discontinued | Female | TT (*1/*1) | 21,347 | 2,489 | 18,290 | 13.6 | *ref* |  |  |  |
| atorvastatin and |  | TC (*1/*5) | 7,538 | 865 | 6,466 | 13.4 | 0.98 | 0.91 | 1.06 | 0.63 |
| simvastatin |  | CC (*5/*5) | 691 | 86 | 589 | 14.6 | 1.06 | 0.86 | 1.32 | 0.55 |
| treatment |  | *Total* | *29,576* | *3,440* | *25,345* | *13.6* |  |  |  |  |
| <1 year after |  |  |  |  |  |  |  |  |  |  |
| first prescription | Male | TT (*1/*1) | 28,609 | 2,333 | 25,743 | 9.1 | *ref* |  |  |  |
|  |  | TC (*1/*5) | 10,056 | 842 | 9,054 | 9.3 | 1.03 | 0.95 | 1.1 | 0.48 |
|  |  | CC (*5/*5) | 947 | 95 | 846 | 11.2 | 1.25 | 1.02 | 1.53 | 0.03 |
|  |  | *Total* | *39,612* | *3,270* | *35,644* | *9.2* |  |  |  |  |
| Discontinued | Female | TT (*1/*1) | 16,950 | 2,987 | 105,435 | 2.8 | *ref* |  |  |  |
| atorvastatin and |  | TC (*1/*5) | 5,977 | 1,081 | 37,192 | 2.9 | 1.03 | 0.96 | 1.1 | 0.41 |
| simvastatin |  | CC (*5/*5) | 542 | 118 | 3,198 | 3.7 | 1.3 | 1.08 | 1.56 | 0.01 |
| treatment |  | *Total* | *23,469* | *4,186* | *145,824* | *2.9* |  |  |  |  |
| >1 years after |  |  |  |  |  |  |  |  |  |  |
| first prescription | Male | TT (*1/*1) | 24,440 | 3,786 | 163,312 | 2.3 | *ref* |  |  |  |
|  |  | TC (*1/*5) | 8,594 | 1,340 | 57,588 | 2.3 | 1 | 0.94 | 1.07 | 0.88 |
|  |  | CC (*5/*5) | 805 | 117 | 5,442 | 2.1 | 0.94 | 0.78 | 1.12 | 0.48 |
|  |  | *Total* | *33,839* | *5,243* | *226,342* | *2.3* |  |  |  |  |
| Discontinued | Female | TT (*1/*1) | 21,347 | 5,476 | 123,725 | 4.4 | *ref* |  |  |  |
| atorvastatin and |  | TC (*1/*5) | 7,538 | 1,946 | 43,657 | 4.5 | 1 | 0.96 | 1.06 | 0.79 |
| simvastatin |  | CC (*5/*5) | 691 | 204 | 3,787 | 5.4 | 1.19 | 1.03 | 1.37 | 0.01 |
| treatment |  | *Total* | *29,576* | *7,626* | *171,169* | *4.5* |  |  |  |  |
| (any time) |  |  |  |  |  |  |  |  |  |  |
|  | Male | TT (*1/*1) | 28,609 | 6,119 | 189,055 | 3.2 | *ref* |  |  |  |
|  |  | TC (*1/*5) | 10,056 | 2,182 | 66,642 | 3.3 | 1.01 | 0.96 | 1.06 | 0.58 |
|  |  | CC (*5/*5) | 947 | 212 | 6,289 | 3.4 | 1.05 | 0.92 | 1.2 | 0.44 |
|  |  | *Total* | *39,612* | *8,513* | *261,986* | *3.2* |  |  |  |  |

Discontinuations = discontinued both simvastatin and atorvastatin prior to data censoring date

### Supplementary Table 3

Switching to another statin from atorvastatin or simvastatin, within 12 months of discontinuation

|  | ***rs4149056 (SLCO1B1 *5) genotype*** | | | | | |
| --- | --- | --- | --- | --- | --- | --- |
|  | **Female** | | | **Male** | | |
|  | **TT (*1/*1)** | **CT (*1/*5)** | **CC (*5/*5)** | **TT (*1/*1)** | **CT (*1/*5)** | **CC (*5/*5)** |
| Pravastatin | 886 | 316 | 26 | 962 | 332 | 34 |
| Fluvastatin | 40 | 13 | 3 | 39 | 16 | 4 |
| Rosuvastatin | 628 | 242 | 25 | 707 | 263 | 21 |
| Cerivastatin | 1 | 0 | 0 | 3 | 0 | 0 |
| *Total* | *1328* | *502* | *50* | *1525* | *540* | *55* |
| Discontinued | 5,476 | 1,946 | 204 | 6,119 | 2,182 | 212 |
| % switched | 24.3 | 25.8 | 24.5 | 24.9 | 24.7 | 25.9 |

Total = number of participants in each sex/genotype group who received a prescription to any another statin within 12 months of their last atorvastatin/simvastatin (some received more than one statin, so the total number of individual statins is greater than the number of people).

### Supplementary Table 4

Last recorded dose of GP-prescribed simvastatin or atorvastatin in the participants who discontinued treatment

|  | ***rs4149056 (SLCO1B1 *5) genotype*** | | | | | |
| --- | --- | --- | --- | --- | --- | --- |
|  | **Female** | | | **Male** | | |
|  | **TT (*1/*1)** | **CT (*1/*5)** | **CC (*5/*5)** | **TT (*1/*1)** | **CT (*1/*5)** | **CC (*5/*5)** |
| Simvastatin |  |  |  |  |  |  |
| 10 mg | 405(10.92) | 136(10.99) | 14(9.21) | 360(8.06) | 109(7.38) | 9(7.09) |
| 20 mg | 1256(33.85) | 424(34.28) | 51(33.55) | 1224(27.39) | 398(26.96) | 39(30.71) |
| 40 mg | 2031(54.74) | 677(54.73) | 85(55.92) | 2858(63.97) | 956(64.77) | 79(62.20) |
| 80 mg | 18(0.49) | 0 | 2(1.32) | 26(0.58) | 13(0.88) | 0 |
| *Total* | *3,710* | *1,237* | *152* | *4,468* | *1,476* | *127* |
| Atorvastatin |  |  |  |  |  |  |
| 10 mg | 1,161(48.93) | 423(50.3) | 37(52.86) | 1075(41.73) | 388(45.33) | 32(33.68) |
| 20 mg | 805 (33.92) | 281(33.41) | 27(38.57) | 929(36.06) | 290(33.88) | 37(38.95) |
| 40 mg | 336 (14.16) | 113(13.4) | 6(8.57) | 454(17.62) | 138(16.12) | 23(24.21) |
| 80 mg | 71(2.99) | 24(2.85) | 0 | 118(4.58) | 40(4.67) | 3(3.16) |
| *Total* | *2,373* | *841* | *70* | *2,576* | *856* | *95* |

### Supplementary Table 5

GP-diagnosed muscle symptoms by SLCO1B1 *5 genotype and sex, stratified by stable treatment period (3 months after first prescription)

| **Model** | **Sex** | **Genotype** | **N cases (%)** | **HR** | **95% CIs** | | ***p*** |
| --- | --- | --- | --- | --- | --- | --- | --- |
| Within | Female |  |  |  |  |  |  |
| 3 months |  | TT (*1/*1) | 74 (0.36) | *ref* |  |  |  |
| of first |  | CT (*1/*5) | 34(0.48) | 1.31 | 0.9 | 2 | 0.18 |
| prescription |  | CC (*5/*5) | 2(0.3) | 0.86 | 0.2 | 3.5 | 0.82 |
|  |  | *Total* | *110* |  |  |  |  |
|  | Male |  |  |  |  |  |  |
|  |  | TT (*1/*1) | 69(0.25) | *ref* |  |  |  |
|  |  | CT (*1/*5) | 25(0.26) | 1.03 | 0.7 | 1.6 | 0.9 |
|  |  | CC (*5/*5) | 2(0.22) | 0.88 | 0.2 | 3.7 | 0.86 |
|  |  | *Total* | *96* |  |  |  |  |
| Greater than | Female |  |  |  |  |  |  |
| 3 months |  | TT (*1/*1) | 584(3.10) | *ref* |  |  |  |
| after first |  | CT (*1/*5) | 244(3.67) | 1.19 | 1 | 1.4 | 0.02 |
| prescription |  | CC (*5/*5) | 20(3.31) | 1.11 | 0.7 | 1.7 | 0.64 |
|  |  | *Total* | *848* |  |  |  |  |
|  | Male |  |  |  |  |  |  |
|  |  | TT (*1/*1) | 736(2.81) | *ref* |  |  |  |
|  |  | CT (*1/*5) | 262(2.85) | 1.01 | 0.9 | 1.2 | 0.85 |
|  |  | CC (*5/*5) | 28(3.22) | 1.16 | 0.8 | 1.7 | 0.43 |
|  |  | *Total* | *1,026* |  |  |  |  |
